# Luminore CopperTouch™ surface coating effectively inactivates SARS-CoV-2, Ebola, and Marburg viruses *in vitro*

**DOI:** 10.1101/2020.07.05.20146043

**Authors:** Emily K. Mantlo, Slobodan Paessler, Alexey Seregin, Alfred Mitchell

## Abstract

We investigated the ability of Luminore CopperTouch™ copper and copper-nickel surfaces to inactivate filoviruses and severe acute respiratory syndrome coronavirus 2 (SARS-CoV-2). For this purpose, we compared viral titers in Vero cells from viral droplets exposed to copper surfaces for 30 min. The copper and copper–nickel surfaces inactivated 99.9% of the viral titer of both Ebola and Marburg viruses. The copper surfaces also inactivated 99% of SARS-CoV-2 titers in 2 hours to close to the limit of detection. These data add Ebolavirus, Marburgvirus, and SARS-CoV-2 (COVID-19) to the list of pathogens that can be inactivated by exposure to copper ions, validating Luminore CopperTouch™ technology (currently the only Environmental Protection Agency-registered cold spray antimicrobial surface technology) as an efficacious, cost-friendly tool to improve infection control in hospitals, long-term care facilities, schools, hotels, buses, trains, airports, and other highly trafficked areas.

## Introduction

Emerging viruses continue to pose a major threat to public health worldwide, as demonstrated by two ongoing outbreaks. Recently, severe acute respiratory syndrome coronavirus 2 (SARS-CoV-2) emerged from a seafood market in Wuhan, China, and quickly spread around the world. This virus is responsible for millions of infections and over 345,523 deaths worldwide and 97,948 in the USA as of May 2020. Viruses from the *Filoviridae* family have caused several outbreaks and epidemics in the past. Ebolavirus caused an outbreak of Ebola virus disease (EVD) in the West African nations of Liberia, Sierra-Leone, Guinea, Nigeria, Senegal, and Mali in 2014–2015, with a current outbreak in the Democratic Republic of the Congo (1, 2). In total, as of May 2020 at least 32,000 people have been infected and approximately 13,600 people have lost their lives due to these outbreaks. Ebolavirus and Marburgvirus, which cause hemorrhagic fevers, present a substantial global threat to public health. Reports from the World Health Organization and the Centers for Disease Control and Prevention (CDC) showed shocking fatality rates from Ebola (∼70%) and Marburg hemorrhagic fever (∼75%) among reported cases during 2015. As of April 2020, a total of 3462 EVD cases, including 3316 confirmed and 145 probable cases, have been reported, with a total of 2279 reported deaths (overall case fatality ratio of 66%). Both Ebolavirus and Marburgvirus originated in Africa, which has experienced the bulk of outbreaks and resultant deaths, but healthcare facilities in Europe and the United States have also reported infected patients.

Many infectious microorganisms can survive for days or weeks after landing on a surface, presenting a tremendous challenge for infection control. In one study, Zaire Ebolavirus and Lake Victoria Marburgvirus were shown to survive in liquid media at titers above detectable limits for up to 46 days, although the number of viable particles decreased over that time (3). When dried on solid surfaces, such as rubber, glass, or plastic, Ebolavirus survives at detectable levels for 5.9 (4) to 14 days (3). SARS-CoV-2 can survive on plastic or stainless steel surfaces for up to 72 h and on copper for 4 h (5). To control the spread of these infections, many guidelines and regulations have been established in hospitals, healthcare and other facilities. Healthcare authorities have instituted routine questioning of patients and visitors regarding their previous geographical location as they enter healthcare facilities (https://www.cdc.gov/coronavirus/2019-ncov/hcp/us-healthcare-facilities.html). Barrier nursing, characterized by isolating patients, wearing protective clothing, and disinfecting surfaces, is key to preventing the spread of both filoviruses and coronaviruses (https://www.cdc.gov/coronavirus/2019-ncov/hcp/infection-control-recommendations.html). To clean healthcare facilities, the CDC requires hospital disinfectants to be effective against certain viruses with genetic structures similar to that of Ebola and coronaviruses. The United States Environmental Protection Agency (EPA) has also created a list of recommended disinfectants that are efficacious against SARS-CoV-2 (https://www.epa.gov/pesticide-registration/list-n-disinfectants-use-against-sars-cov-2-covid-19).

Commensurate with the evaluation of these preventive efforts, attention has turned to the feasibility of self-sanitizing surfaces. Copper exhibits a self-sanitizing characteristic, but its weight and bulk have limited its use. To circumvent these challenges, antimicrobial copper coatings have been developed. Luminore CopperTouch™ is an EPA-registered antimicrobial copper surface. Copper and copper–nickel formulations are currently registered and available for application in hospitals, doctor’s offices, airports, buses, trains, and other heavily trafficked areas. These antimicrobial coatings greatly expand the possibility of using copper within the transportation and healthcare settings, particularly for high-touch surfaces, to reduce the spread of infection. Employing these coatings to prevent infection is preferable to attempts at treatment or curing the disease after infection occurs.

Several studies have demonstrated the effect of copper in reducing the bacterial burden in hospitals and healthcare facilities. Salgado *et al*. reported a significant reduction in hospital-acquired infections and/or methicillin-resistant *Staphylococcus aureus* or vancomycin-resistant *Enterococcus* colonization for patients that received treatment in intensive care unit (ICU) rooms with copper alloy surfaces compared with standard ICU rooms (6). Viruses are susceptible to copper surfaces as well, including influenza A virus (7) and norovirus (8, 9), both of which are inactivated by copper. Solid copper surfaces have also been shown to inactivate SARS-CoV-2 (5).

Because Ebolavirus and Marburgvirus are considered biosafety level (BSL)-4 agents and SARS-CoV-2 is considered a biosafety level (BSL)-3 agent, the capacity to study these viruses is limited to specially equipped, government-registered laboratories. We investigated the effect of copper and copper–nickel surfaces on the viability of Ebolavirus and Marburgvirus. We found that these surfaces begin to inactivate the viruses within 15 min, reducing the viral load to 1%? 3% of the initial titer within 30 min. These findings add Ebolavirus and Marburgvirus to the growing list of infectious microorganisms that are inactivated upon copper exposure. We also investigated the effect of Luminore CopperTouch™ copper-coated surfaces on SARS-CoV-2 and observed a significant drop (99%) in viral titer after 2 hours. These findings lend strong support to the use of Luminore CopperTouch™ and other copper surfaces in hospitals, buses, trains, and public areas to reduce the spread of viral infections, with substantial implications for public health.

## Results

Copper-coated surfaces inhibit the growth of Gram-positive and Gram-negative bacteria, as well as influenza virus, norovirus, and human immunodeficiency virus (HIV). To determine whether copper and copper–nickel surfaces can effectively reduce the viral load of Ebolavirus and Marburgvirus after exposure, 10-μl droplets of viral suspension were exposed to Luminore CopperTouch™ surfaces for 30 min at room temperature. Marburgvirus was also tested in 10 1-μl droplets. We measured the viral load using a standard plaque assay in Vero E6 cells. We observed a 99.9% reduction in viral titer on copper surfaces for MARV and EBOV relative to the sham surfaces after 30 min (Fig. 1), approaching the limits of assay detection. These data suggest that the CopperTouch surfaces can effectively inactivate EBOV and MARV particles.

**Figure 1.**
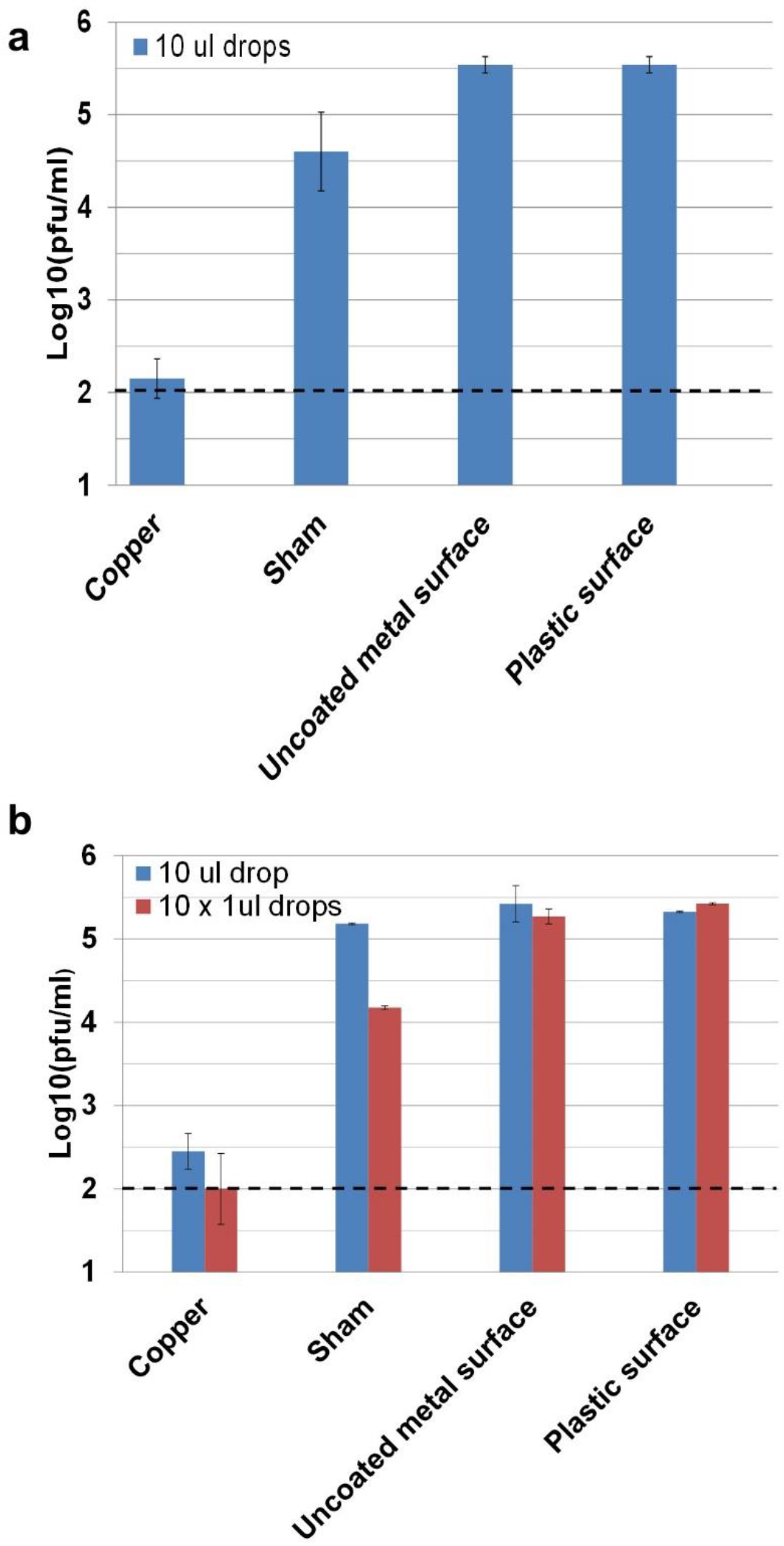
Ebolavirus (Zaire) and Marburg virus (Angola) are inactivated on copper and copper–nickel surfaces. (A) EBOV (10-μl drop) and (B) MARV (10-μl drop or 10 × 1-μl drops) viruses were exposed to the indicated surfaces for 30 min and were then used to infect Vero E6 cells. Standard plaque dilution assays were utilized to calculate the viral titer. The dashed horizontal lines indicate the limit of detection.

To determine how rapidly copper surfaces can inactivate viral particles, we measured EBOV viral titers using plaque assays on Vero cells after viral suspensions had been exposed to surfaces for 1, 15, or 30 min (Fig. 2). At 1 and 15 min, the viral titers decreased by approximately 0.5 log-fold relative to the sham surface. After 30 min, the copper surface had reduced the viral load approximately1.5 log-fold, corresponding to a 97% reduction in viral titers. The copper–nickel surface displayed a decrease of 2.3 log-fold, corresponding to a reduction of more than 99% in viral titers. These data suggest that viral inactivation occurs within 15–30 min and that the viral load is reduced by nearly 99% within that time period.

**Figure 2.**
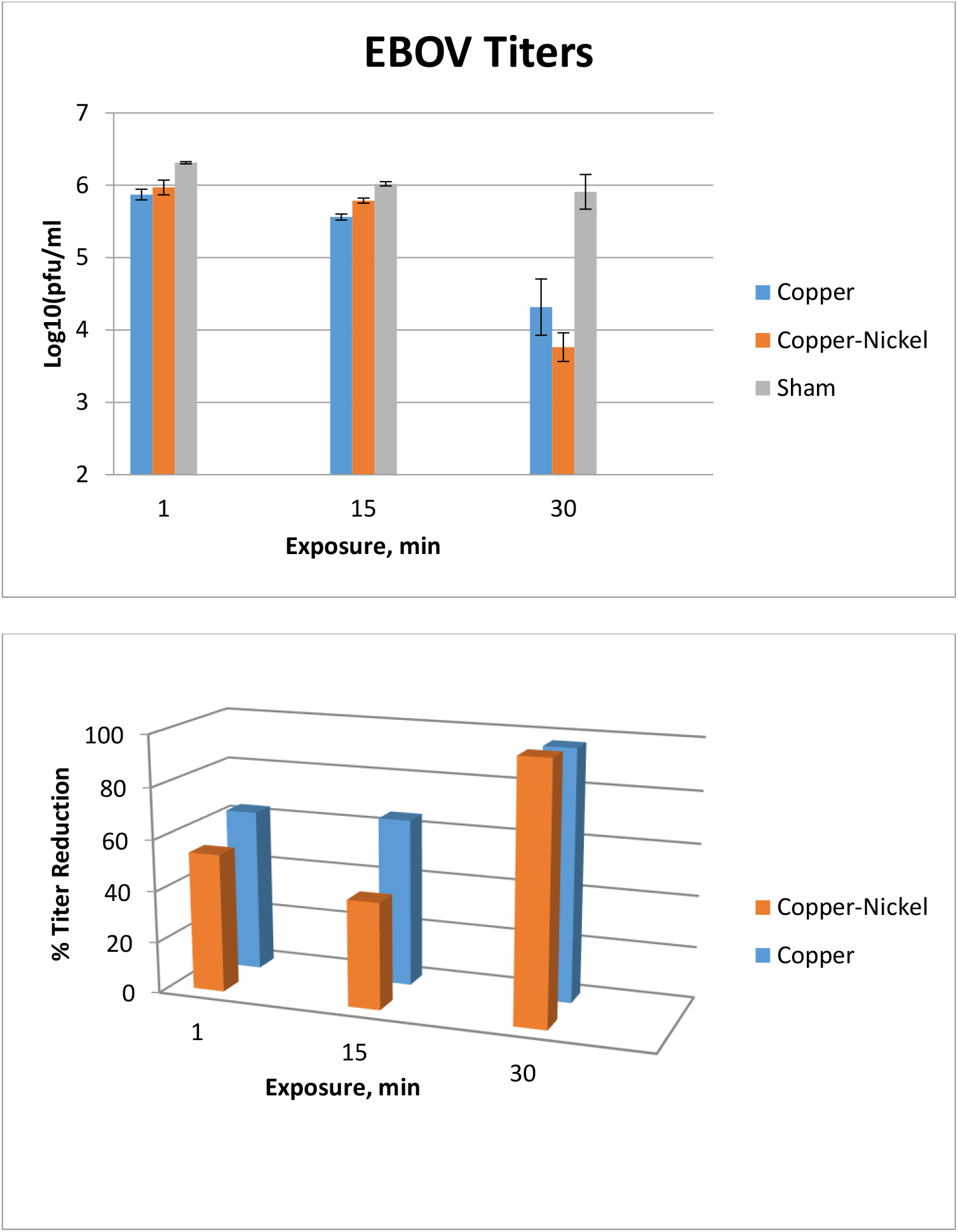
Ebolavirus is inactivated within 30 min of exposure to copper and copper–nickel surfaces. Suspensions of Zaire Ebolavirus (EBOV) were exposed to each surface for the indicated amount of time, and the exposed viruses were then used to infect Vero cells. Standard plaque dilution assays were utilized to calculate the viral titer. Copper (triangle), copper–nickel (square), and sham (diamond) surfaces were compared for up to 30 min. Data are reported as the calculated average titers for treatments performed in duplicate to minimize BSL-4-level work.

Finally, we assessed the ability of copper surfaces to inactivate SARS-CoV-2 particles. We exposed copper, metal, and plastic surfaces to SARS-CoV-2 for 2, 4, or 8 hours and measured viral titers via TCID_50_. Regardless of the incubation time, the copper-coated surfaces reduced the viral titers greater than 2-fold, approaching the limit of detection (Fig. 3). This value corresponds to a reduction of more than 99% in viral titers.

**Figure 3:**
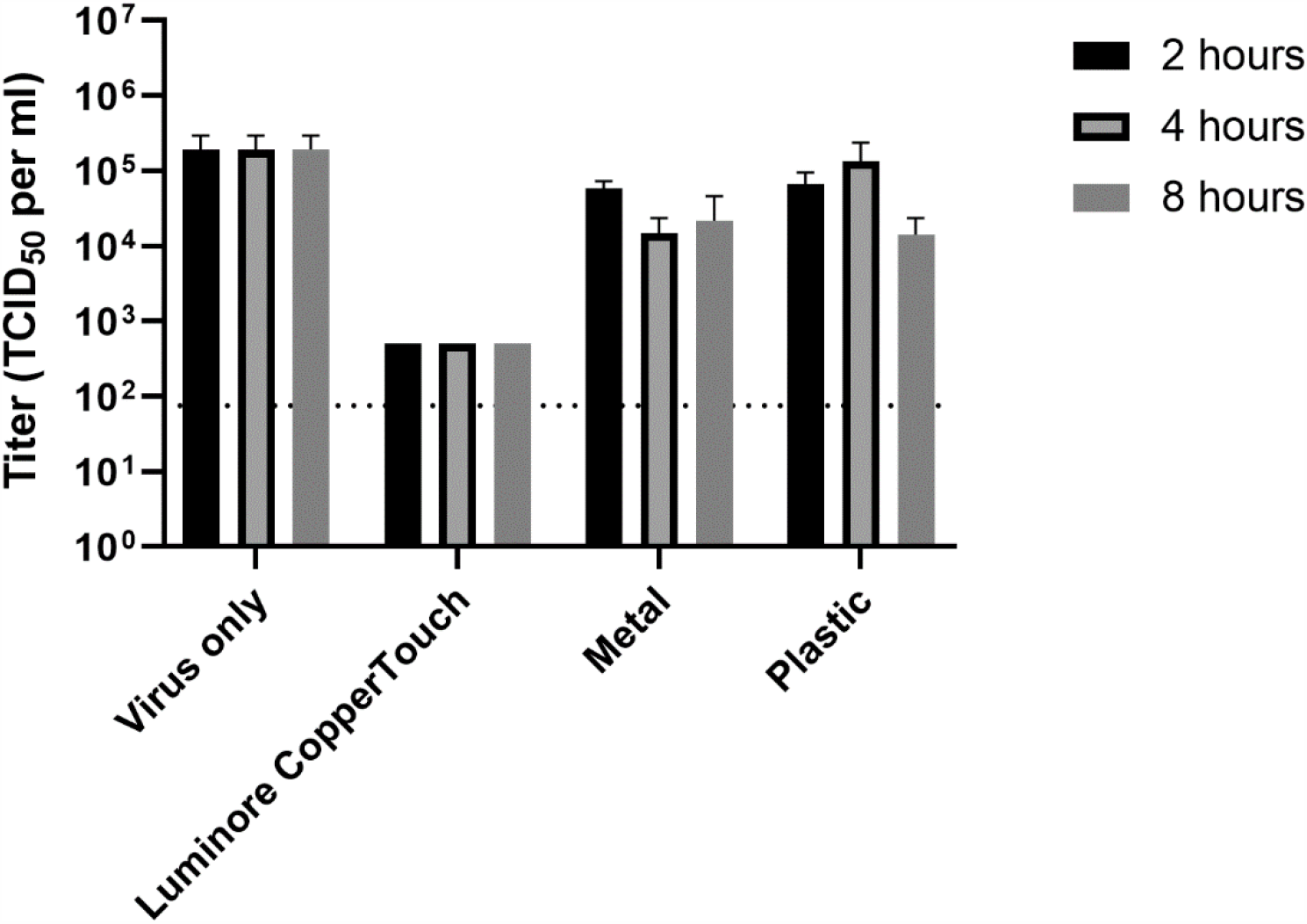
SARS-CoV-2 is inactivated within 2 hours of exposure to copper-coated surfaces. One 10-µl drop of SARS-CoV-2 was added to the indicated surfaces for the denoted amount of time. The samples were collected with media and were used to immediately infect Vero cells for calculating the viral titer via TCID_50_. The data shown represent the average titers for experiments performed in triplicate. The error bars display the standard distribution, and the dotted line indicates the limit of detection.

## Discussion

In this study, SARS-CoV-2 titers were reduced by more than 99% after 2 hours. Both MARV and EBOV viral titers were reduced by more than 99% on copper surfaces within 30 min. Previous research has demonstrated that EBOV can persist on dry surfaces for 1−2 weeks (3, 4). By reducing the persistence rate, Luminore CopperTouch™ copper surfaces can shorten the viral particle survival time by up to 336-fold compared to plastic or metal surfaces. Copper-coated surfaces can also significantly reduce SARS-CoV-2 survival in comparison to plastic or metal surfaces. These findings have far-reaching implications for patient welfare and healthcare costs. Of immediate interest is the use of these coatings in healthcare, public transportation, and other settings to prevent the spread of viruses. By mitigating the presence, growth, and spread of such pathogens on frequently touched surfaces, copper coatings can play an important role in infection control and disease prevention.

Research on the antiviral and antibacterial properties of copper alloys and other metals (e.g., silver, iron) has increased tremendously over the past five years. Researchers have turned to self-sanitizing surfaces as a supplementary infection control strategy, given the challenges of vaccine and antibiotic development. In addition to copper, surfaces containing silver and zinc also exhibit antimicrobial properties. Sehmi *et al*. developed a method for encapsulating silicone and polyurethane with copper, which both exhibited potent bacterial activity (10). By comparing thick (100-μm) and thin (25-μm) rolled copper plates, researchers have noted that thin-rolled sheets are rougher in texture than thick-rolled sheets. However, the data comparing their relative effectiveness are contradictory. While Zeiger *et al*. demonstrated that smoother copper surfaces have more antibacterial activity than rough surfaces (11), Yousuf *et al*. showed that thin-rolled sheets have more potent activity and attributed this effect to the increased surface area of the rough surfaces (12). More recently, a hybrid coating containing silver, copper, and zinc cations was found to significantly reduce viral titers for HIV-1, human herpesvirus 1, dengue virus type 2, and influenza H1N1 virus (13). With these advancing breakthroughs in the design of self-sanitizing surfaces, our finding that Luminore CopperTouch™ copper surfaces inactivate viruses of critical public health concern adds to the enthusiasm for copper surfaces in hospitals and public spaces to aid in infection control.

The mechanism by which copper inactivates microorganisms is not completely understood. Winstein *et al*. demonstrated that copper can donate and accept single electrons, which produces reactive oxygen species and free radicals, causing cell death. In another study, Warnes *et al*. reported that copper surfaces inhibit the transfer of DNA (i.e., plasmids harboring antibiotic resistance mechanisms) between multidrug-resistant bacteria (14). The ability of copper to inhibit bacteria involves the rapid degradation of genomic and plasmid DNA. This study further showed that DNA degrades rapidly on copper surfaces (14). Thus, it is plausible to hypothesize that the genomes of these viruses are disrupted by the free radicals produced upon contact with copper ions.

In conclusion, we have shown that the copper and copper–nickel alloys produced by Luminore CopperTouch™ have a potent antiviral effect against Ebolavirus, Marburgvirus, and COVID-19. These data have significant implications for infection control, particularly during times of regional epidemics, for hospitals and other highly trafficked areas, such as doctor’s offices, prisons, airports, buses, trains, and schools. Additionally, these data could influence strategies for epidemic relief for both ongoing and future epidemics, such as the current SARS-CoV-2 pandemic and the ongoing EBOV epidemic in the Democratic Republic of Congo, by supporting the use of copper coatings on equipment sent to highly affected areas.

### Significance and Impact

In this study, data regarding the survival of Ebola, Marburg, and SARS-CoV-2 viruses on copper (Luminore CopperTouch™) are presented for the first time. These data will aid in the implementation of strategies to help prevent infection and transmission of disease.

## Materials and Methods

### Viral strains and propagation

Filoviruses were maintained in a government-registered BSL-4 facility at the University of Texas Medical Branch in Galveston, TX, in compliance with all regulations therein. Zaire ebolavirus (EBOV; Stock) and Angola Marburg virus (MARV; Stock) were utilized, and SARS-CoV-2 (USA-WA1/2020) was obtained from the World Reference Center for Emerging Viruses and Arboviruses (WRCEVA). All experiments with SARS-CoV-2 were conducted in approved BSL-3 facilities at the University of Texas Medical Branch.

### Cell culture

Vero or Vero E6 cells were cultured and maintained in Eagle’s minimum essential medium (MEM) with Earle’s BSS (balance salt solution) and L-glutamine (EMEM) or Dulbecco’s modified Eagle medium. The media were supplemented with fetal bovine serum (FBS; 1%–2%) and 100 μg/ml penicillin/streptomycin (pen/strep) solution. This same medium was used as dilution medium for plaque assays. The cells were incubated at 37±2°C with 5% CO_2_.

### Surfaces

LuminOre(r) is a unique patented cold spray liquid metal. It is a polymetal alloy, not a paint. To the best of our knowledge, LuminOre is the only company that has ever been able to blend cold alloy metals and polymers together to form this type of homogeneous metal matrix. This liquid metal is then cold sprayed onto any substrate to form a solid metal surface. The Luminore CopperTouch™ copper surface is 85% copper, and the copper–nickel alloy is a proprietary blend containing 62.5% copper. For surfaces, 1-mm thick, 1 × 1-cm coupons were employed.

### Filovirus surface exposure

The viral load consisted of 10^5^ plaque-forming units (pfu) and viral suspensions maintained in growth medium solution (DMEM with 1x L-glutamine, 1x pen/strep, 1% MEM vitamins, and 10% FBS). Vero E6 cells were grown to 70%−80% confluence in 6- or 12-well plates. Ten 1-μl drops (MARV) or one 10-μl drop (MARV and EBOV) of viral suspensions were dispensed on either copper, copper–nickel, or sham coupons (as described above) or on uncoated metallic or plastic surfaces as controls, with a subsequent 30-min incubation at room temperature. The drops were then collected with 100 μl of fresh dilution medium and titrated on Vero E6 cells using a standard plaque assay technique (0.5% agarose overlay) (15). Inoculated Vero E6 cells were incubated at 37±2°C with 5% CO_2_ for 7 (MARV) or 10 days (EBOV). The titrations were subsequently fixed with formalin. To enumerate the plaques, the wells were stained with neutral red or crystal violet solutions using standard procedures. To minimize the extent of work performed in the BSL-4 laboratory and the production of waste, experiments were conducted in duplicate.

### Coronavirus surface exposure

Vero cells were grown to 85%–95% confluence in 96-well plates. One 10-µl drop of SARS-CoV-2 stock (5 x 10^5^ TCID_50_/mL [Tissue Culture Infectious Dose 50%/mL]) was added to copper-coated or uncoated metallic or plastic surfaces as controls and were incubated for the indicated duration at room temperature. These experiments were conducted in triplicate. The drops were collected with 90 µl of fresh dilution medium and titrated on Vero cells using a TCID_50_ assay.

## Data Availability

Extracted data are available on request to the corresponding author.

## Authors’ Contributions

SP and EKM conducted the experiments, AS assisted in the laboratory procedures and data collection, and AM led the project.

## Acknowledgements

We thank Drs. Kenneth Plante (The World Reference Center for Emerging Viruses and Arboviruses, UTMB) and Natalie Thornburg (U.S. Centers for Disease Control and Prevention) for providing the SARS-CoV-2 stock virus. We thank Tom Valente for support, technical assistance, and funding. EKM was supported by NIH T32 training grant AI060549.

